# The pathogenic p.R391G ABCC6 displays incomplete penetrance implying the necessity of an interacting partner for the development of pseudoxanthoma elasticum

**DOI:** 10.1101/2020.11.26.20236489

**Authors:** Flora Szeri, Agnes Miko, Nastassia Navasiolava, Ambrus Kaposi, Shana Verschuere, Qiaoli Li, Sharon F. Terry, Federica Boraldi, Jouni Uitto, Koen van de Wetering, Ludovic Martin, Daniela Quaglino, Olivier M. Vanakker, Kalman Tory, Tamas Aranyi

## Abstract

ABCC6 promotes the efflux of ATP from hepatocytes to the bloodstream. ATP is then cleaved to AMP and pyrophosphate, a major inhibitor of ectopic calcification. Pathogenic variants of ABCC6 cause pseudoxanthoma elasticum, a recessive ectopic calcification disease of highly variable severity. One of the mechanisms influencing the heterogeneity of a disorder is the penetrance of pathogenic variants. Penetrance shows the proportion of carriers developing the phenotype; hence incomplete penetrance indicates that the disease does not necessarily develop in the presence of specific variants. Here, we investigated whether incomplete penetrance contributes to the heterogeneity of pseudoxanthoma elasticum. By integrating the clinical and genetic data of 589 patients, we created the largest European cohort. Based on allele frequencies compared to a reference cohort, we identified two incomplete penetrant variants, p.V787I and p.R391G, 6.5% and 2% penetrance, respectively. The characterization of the p.R391G variant suggested unaltered severity of the clinical phenotype. Based on our biochemical and localization studies, we hypothesize that the variant becomes deleterious only if an interacting partner is mutated simultaneously. Our data reveal the potential existence of the first interacting partner of ABCC6. Our data are also important for genetic counseling, as they suggest lower disease heritability of some variants.

## Introduction

*ABCC6* encodes a transmembrane ATP-binding cassette protein primarily expressed in the liver (Favre et al. 2017). The protein promotes the efflux of intracellular ATP to the bloodstream (Jansen et al. 2014). ATP is subsequently cleaved by ENPP1 to AMP and pyrophosphate (PPi), a major inhibitor of ectopic calcification (Back et al. 2018; Dedinszki et al. 2017). Bi-allelic loss-of-function (LOF) pathogenic variants of ABCC6 lead to pseudoxanthoma elasticum (PXE) (Ringpfeil et al. 2000; Le Saux et al. 2000), a slowly progressing disease. PXE is characterized by soft tissue calcification leading to dermatologic, ocular, and cardiovascular manifestations including vascular calcification, intermittent claudication, and higher incidence of cardiovascular events (De Vilder et al. 2018; Köblös et al. 2010). The prevalence of PXE is ∼1/25,000-50,000 (De Vilder et al. 2018; Köblös et al. 2010) and over 350 pathogenic variants were reported in the several thousands of patients recruited worldwide mainly by the PXE International patient organization and the European PXE reference centers (Legrand et al. 2017; Larusso, Ringpfeil, and Uitto 2010; Moitra et al. 2017; Boraldi et al. 2021).

It is well-known for several monogenic diseases that not all individuals harboring a disease-causing genotype do develop the clinical phenotype (Moore et al. 1993; Ahluwalia et al. 2009; Venturini et al. 2012), a phenomenon called incomplete penetrance (IP). Some mutations can have a penetrance below 5%, which makes genetic counseling challenging (Minikel et al. 2016; Mikó et al. 2021). Although IP is a well-known phenomenon in autosomal dominant diseases where the increased expression of the wild-type allele can counterbalance the mutated allele (Ahluwalia et al. 2009; Venturini et al. 2012), it was more recently also detected in recessive disorders in large pedigrees (Israel et al. 2017), found in cystic fibrosis (*CFTR*) (Mikó et al. 2021), and NPHS2-associated Steroid-Resistant Nephrotic Syndrome (Tory et al. 2014).

IP can be assessed by the characterization of patient cohorts requiring reference populations now available as a result of the worldwide sequencing efforts (Wright et al. 2019). Using a recently developed algorithm (Mikó et al. 2021), which compares the allele frequencies (AF) in a patient cohort to a reference population, the penetrance of a pathogenic variant in the cohort can be assessed. If a sequence variant in a patient cohort of a recessive disease is not significantly more frequent (i.e., not significantly enriched) relative to the general population, it is considered to be pathogenic erroneously. Nevertheless, it is possible that some sequence variants are significantly enriched in the patient cohort, albeit they are significantly less enriched than the other pathogenic alleles. These alleles are disease-causing but exhibit IP. Individuals harboring such an incomplete penetrant allele can therefore manifest the disease but may also remain completely free of the symptoms (Mikó et al. 2021).

In this study we investigated whether pathogenic PXE alleles exhibit incomplete penetrance. We studied phenotypically and genotypically characterized patient cohorts to shed more light on the mechanism of the disease and improve genetic counseling in PXE.

## Results

### Characteristics of the largest European PXE patient cohort

We first compiled an anonymized cohort of clinically (Phenodex score (Pfendner et al. 2007) plus age) and molecularly diagnosed PXE patients. Altogether 589 probands with European ancestry from three countries (Belgium, France, and Italy (Vanakker et al. 2008; Boraldi et al. 2021)) were collected (Supplementary Table 1A). Only patients of known ethnic origin, familial relationship, and with two identified *ABCC6* variants considered to be disease-causing at the time of diagnosis were included in our cohort, which contained 216 different variants including missense, non-sense, frameshift variants and large deletions (Figure 1A and Supplementary Table 1B). (Uitto, Li, and Jiang 2010; Nitschke et al. 2012; Omarjee et al. 2019).

**Figure 1.**
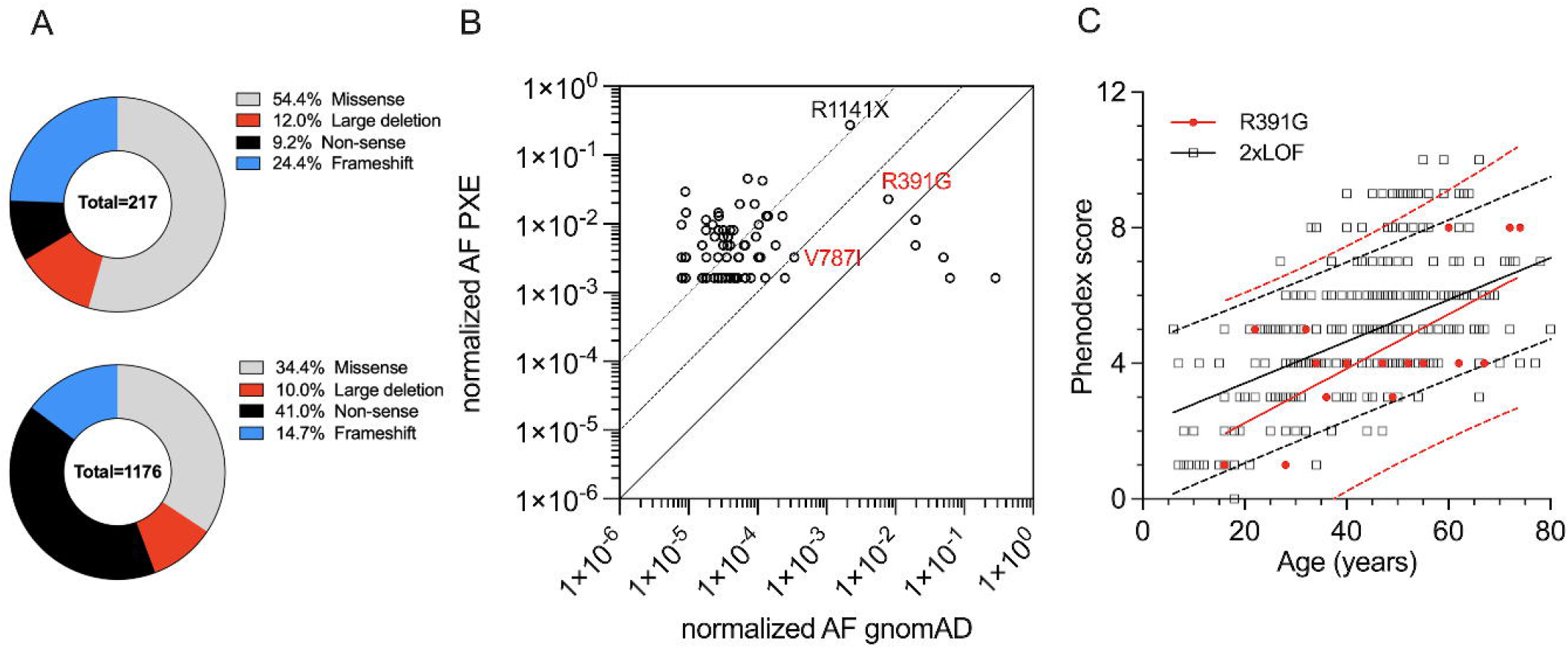
Enrichment and severity of p.R391G pathogenic variant in the European PXE patient population. A) Distribution of frequency of mutation types in the European patient cohort of 589 individuals. The upper panel depicts the ratio of each allele variant falling into the four categories. The lower panel shows the actual allele frequency of variants in the cohort. B) Enrichment of sequence variants in the patient population relative to the European non-Finnish population of gnomAD database. Each dot represents the AF of an individual sequence variant in the 2 databases in a logarithmic scale. Solid, dashed, and punctuated lines represent 1x, 10x, and 100x enrichments, respectively. The incomplete penetrant p.R391G and p.V787I and the most frequent p.R1141* are also indicated. The sequence variants not represented on this logarithmic plot because they are absent from the gnomAD database are shown is Supplementary table 1C) Relationship between Phenodex score and age. Open squares represent the sum of Phenodex scores of individual patients with 2 truncating pathogenic variants. Red circles represent patients with one p.R391G pathogenic variant. Black and red lines represent linear regression of the patients with 2xLOF and p.R391G, respectively. Dotted lines matching the line colors show 95% confidence intervals for the linear regression. (Y_2xLOF_ = (0.062± 0.007)*X + 2.182± 0.323; Y_R391G_ = (0.080±0.024) *X + 0.633±1.175; R^2^_2xLOF_ =0.551, R^2^_R391G_ =0.452)

### Frequency of ABCC6 sequence variants in PXE patients and the general population

Our cohort was first filtered to prevent the overrepresentation of certain alleles by including several members of the same family. The recently developed algorithm (Mikó et al. 2021) excluded patients with indels >50bp as the database of the reference population (the gnomAD database; https://gnomad.broadinstitute.org) doesn’t contain such large deletions. As the result of the stringent pipeline, only 616 alleles remained from the initial 589 patients.

Subsequently, an enrichment analysis was performed by comparing the AF of the remaining variants of the patient cohort and the European non-Finnish population of the gnomAD database (Karczewski et al. 2020) (Figure 1B and Supplementary Table 1C). Most of the 616 alleles in the patient cohort are present as a single cluster, far above the 1 times enrichment threshold, demonstrating their full penetrance and probable pathogenicity. In this cluster p.R518* (n=28), p.R518Q (n=26), p.Q378* (n=18), c.G3736-1G>A (n=12), p.R1138W (n=12) were represented by at least 10 alleles (AF >1.5% in the cohort). Importantly, the majority of the previously reported pathogenic variants were indeed significantly enriched in the patient cohort relative to the general population.

Strikingly, p.R1268Q, p.R265G, c.3507-3C>T, p.L946I, c.346-6G>A were not enriched in the patient population (below the 1 times enrichment threshold). This means that these variants are more frequent in the general population than in the patient population, thus most likely represent benign, non-pathogenic variants that were probably mistakenly identified previously as disease-causing. We excluded them from further investigations.

Two additional outliers of the cluster of the pathogenic variants were identified: p.R1141*, which had a high AF both in the general and the patient population (125-fold enrichment) accounting for almost 1/3 of all PXE-causing mutations and p.R391G, which had an unexpectedly high AF in the general population (0.78%) and showed a small but significant enrichment amongst patients (2.89-fold enrichment; p=5×10^-4^). This relatively low enrichment raised the possibility that this variant was incompletely penetrant.

### Identification of incompletely penetrant ABCC6 variants

In the second part of the analysis, we performed the IP test on the significantly enriched alleles. The algorithm compared the AF of each variant to the combined AF of fully penetrant complete LOF variants, where no full length ABCC6 is present. We considered a pathogenic variant to be incompletely penetrant if the result of the comparison was lower than 30%, a threshold set arbitrarily (Mikó et al. 2021). We thus identified two variants as incompletely penetrant: p.V787I and p.R391G. The p.V787I, despite of its low frequency, was enriched significantly in the PXE cohort relative to the general population (9.53-fold; p=0.02). Nevertheless, this enrichment was much less prominent than that of the LOF pathogenic variants (Supplementary Table 1D), demonstrating potential IP of the p.V787I (6.5%; p<5.1*10^-8^), suggesting that in >90% of the cases the homozygous or compound heterozygous presence of this variant does not lead to the development of PXE.

In the followings we focused on p.R391G, the second most frequent missense mutation in our patient cohort (carried by 17/589 patients), for which we calculated an ∼ 2% penetrance (p<2.6*10^-118^). This suggests that only 2 p.R391G heterozygous carriers would develop PXE out of 100 individuals when carrying a completely penetrant pathogenic variant on the other allele.

The international US PXE cohort, although similar in size (478 patients) as the European cohort, lacked details about familial relationship and ethnicity, and was therefore only used to validate the IP of the p.R391G pathogenic variant, instead of combining it with the European cohort. The US cohort contained eleven patients that were compound heterozygous for the p.R391G. In five patients the p.R391G variant the only detected pathogenic ABCC6 variant. Comparing both patient cohorts revealed very similar AF and IP when matched with the gnomAD database.

To assess the severity of the PXE phenotype in the presence of the p.R391G variant, we compared the Phenodex scores of patients with those having two LOF alleles (2xLOF) in the European cohort. Here, we also included patients with large deletions not represented in the gnomAD database and all members of the same family to increase the power of this comparison. Plotting the Phenodex scores as a function of age (Figure 1C), as expected, indicated disease worsened with age. Linear regression analysis showed that the fitted lines were not significantly different between patients with the p.R391G variant and patients with (2xLOF) variants implying similar disease severity and progression.

### Genetic mechanisms of IP

In the followings, we tested whether the observed IP of p.R391G could be explained by an effect on splicing, which frequently causes reduced penetrance (Rave-Harel et al. 1997; Cogan et al. 2012) or by its obligate association with another pathogenic variant(s) in the *ABCC6* gene. Although the variant causing p.R391G is close to the 3’ end of the exon 9 (n-5 bp), splice site prediction indicated no altered splicing (ASSP) (Wang and Marín 2006). This could not be further confirmed by in vivo gene expression analysis due to the absence of patient tissue samples, as the gene is primarily expressed in the liver.

Next, we tested the potential association of p.R391G with any specific pathogenic variant of *ABCC6* on the other allele. We found that from the initial 17 European patients with p.R391G only 3 had missense pathogenic variants on the other allele. All others were associated with LOF pathogenic variants. This result ruled out the specific association of a missense variant on the 2^nd^ allele in PXE patients with p.R391G. Next, we investigated whether p.R391G is more prone to be associated with truncating than missense variants. In the original European patient cohort, there were almost 2 times more LOF pathogenic variants compared to missense pathogenic variants. In the filtered table of 616 alleles 58% of the pathogenic variants were truncating (345 vs 247 after exclusion of the 14 remaining patients with p.R391G alleles). The association of LOF variants with the incomplete penetrant allele (11/14) was therefore not significantly increased relative to the proportion of LOF pathogenic variants in the patient cohort (p=0.17). Altogether we did not observe any pathogenic variant associated from the second allele with the incomplete penetrant allele.

Next, we postulated that the R391 residue is involved in a pivotal intramolecular interaction. We assume that this interaction is altered by the substitution of arginine by glycine only if another residue is also mutated in ABCC6. This hypothesis was supported by the analysis of the conservation of the R391 residue and homology models of ABCC6. The R391 and flanking residues encoded by exon 9 and 10 are well conserved throughout evolution (Figure 2A) suggesting that they are important for the protein function (Clustalw) (Thompson, Higgins, and Gibson 1994). Furthermore, R391 is also conserved in other members of the ABCC family (ABCC1, ABCC3, and ABCC4), while it is replaced by the similarly charged lysine in ABCC2 and ABCC7 (CFTR) (Ran, Zheng, and Thibodeau 2018) (Figure 2 B). The position of the R391 amino acid residue according to the most recent homology model built on the crystal structure of the bovine ABCC1 protein indicates that the residue is in the 4^th^ intracellular loop at the cytoplasmic surface of ABCC6 (Issa, Tysoe, and Caswell 2020). This model also shows that R391 and the entire loop has a different localization in the inward and the outward facing conformations of the transporter, suggesting a potential role of this domain in the conformational switch necessary for the function of ABCC6. Furthermore, the model revealed potential intramolecular interactions of R391 with V249, E253, V1147, and N1151. Based on these findings we tested whether there was an association of the p.R391G pathogenic variant with any of the coding SNPs of the *ABCC6* gene in the patients. This analysis did not reveal any R391G associated SNPs in *ABCC6* either among the European or the US patients. We, therefore, concluded that the disease-causing mechanism of the p.R391G pathogenic variant is not based on the alteration of an intra-protein interaction due to the presence of a simultaneous ABCC6 SNP affecting a second amino acid in the protein.

**Figure 2.**
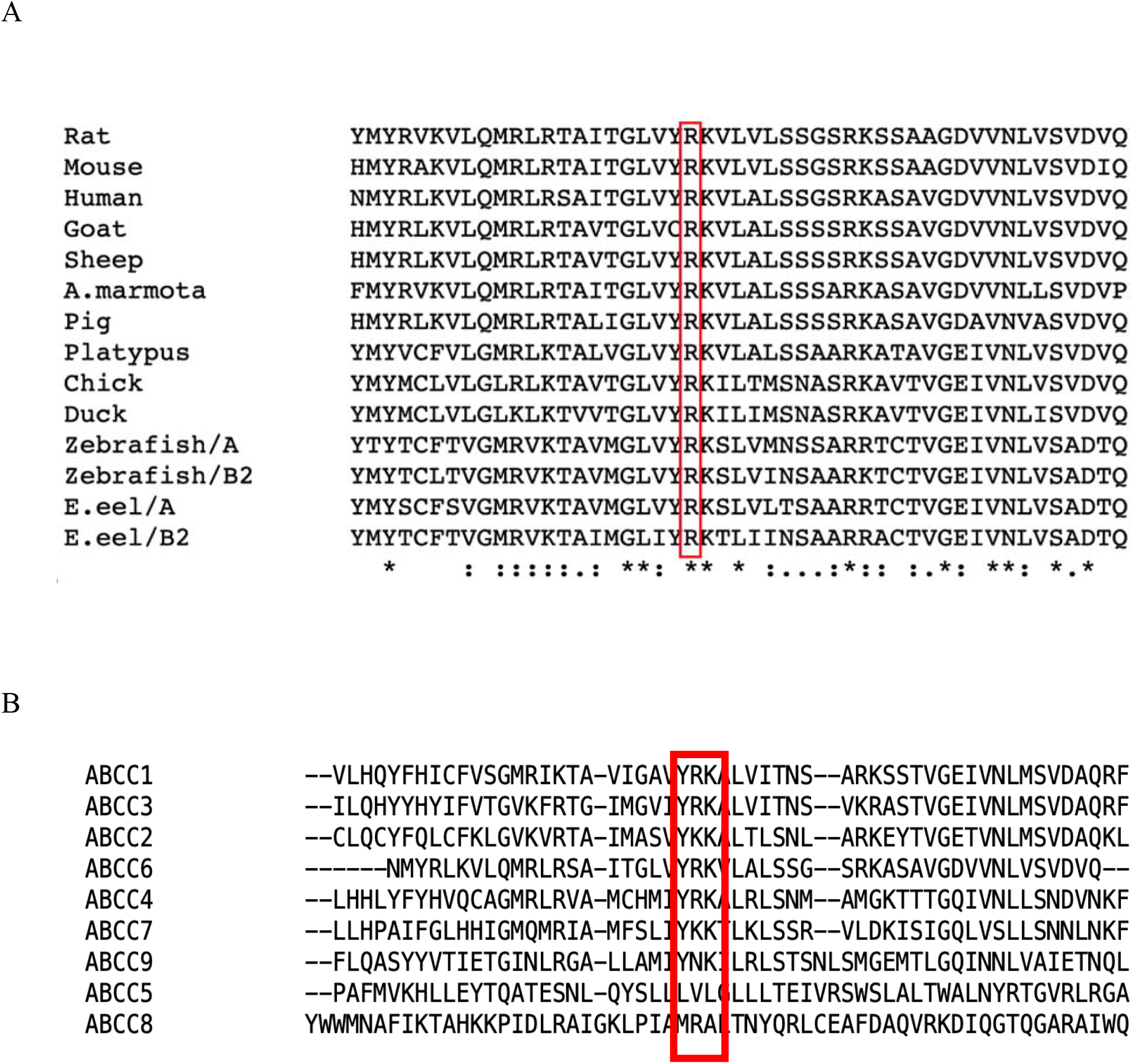
A) Alignment of the sequence flanking the R391 amino acid residue in various vertebrates. The region adjacent to R391 of ABCC6 is highly conserved throughout evolution. The red rectangle indicates the alignment of R391, which is identical in all species analyzed from human to zebrafish and electric eel. Of note, in the two latter species there are two ABCC6 genes. B) Alignment of the sequence flanking the ABCC6 R391 amino acid residue in various members of the ABCC family in human. The red rectangle indicates the R391 and its two neighbors.

### Functional characterization of the incomplete penetrant variant

We hypothesized that due to its low penetrance (∼2%) the p.R391G ABCC6 would behave similarly to wild-type ABCC6 protein in *in vitro* experiments. For these critical experiments we overexpressed the rat ABCC6 protein, previously shown to be significantly more active than the human ortholog with the region of interest being fully conserved (Jansen et al. 2014; Jansen et al. 2013), in HEK293 cells. Of note, the residue homologous to the human R391 in rat is R389 (Figure 2A). The expression levels of the proteins, compared by Western-blot analysis in different clones, were similar (Figure 3A), as well as the plasma membrane localization of the overexpressed wild-type and mutant ABCC6 (clone A2) (Figure 3B). This suggested that the variant did not alter the localization of the protein confirming our expectations and further supporting the hypothesis that the identified pathogenic variant is indeed incompletely penetrant.

**Figure 3.**
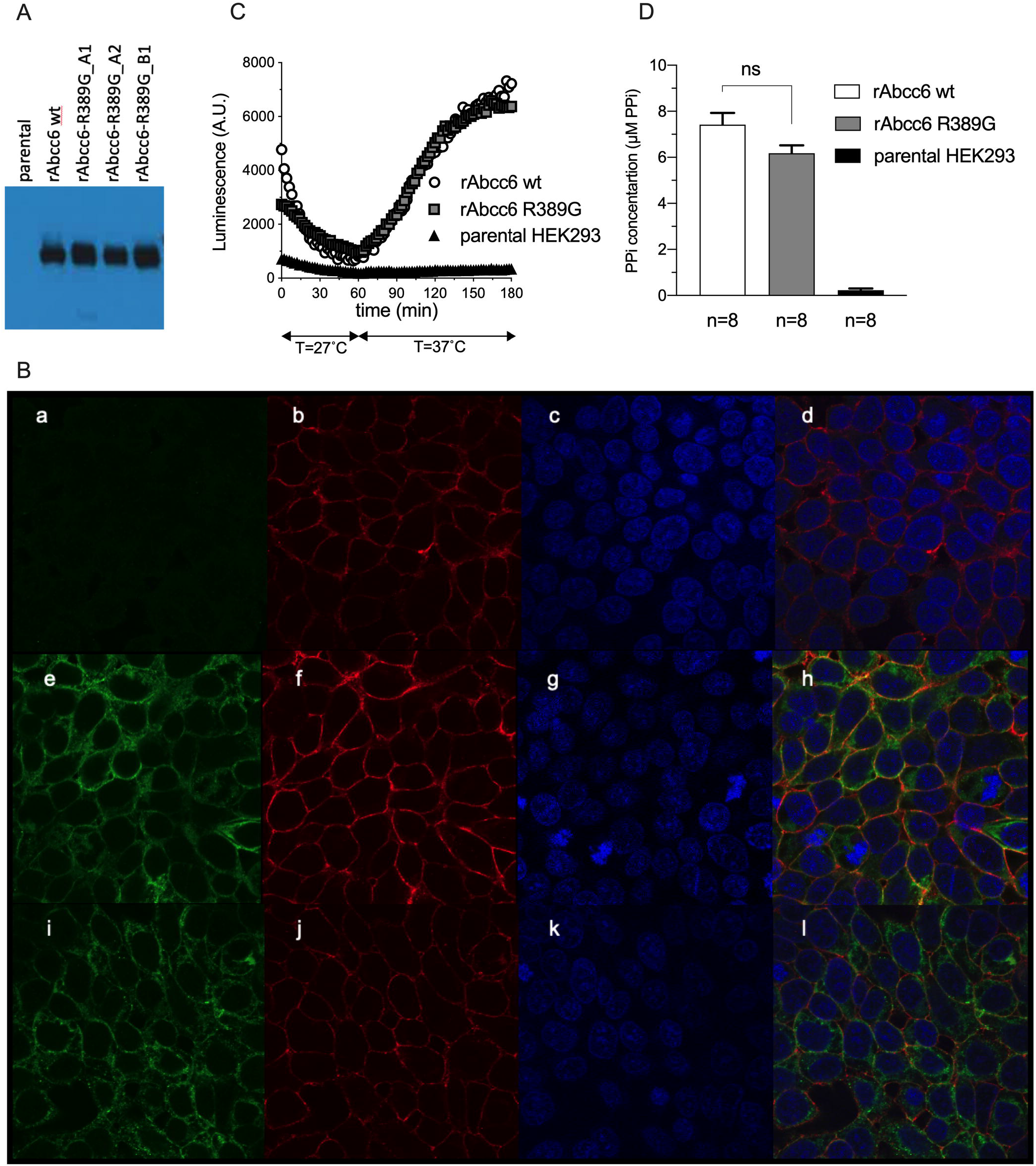
In vitro characterization of the human ABCC6 p.R391G equivalent rat ABCC6 R389G pathogenic variant. A) Overexpression of the wild-type and rat p.R389G mutant ABCC6 in HEK293 cells. 5µg protein of total cell lysates were shown on a Western-blot. The A2 clone of rat ABCC6 R389G mutant was selected for further analysis based on its similar expression level compared to that of the wild type rat ABCC6. B) Representative images showing the subcellular localization of the overexpressed wild-type and p.R389G mutant ABCC6 in HEK293 cells by confocal microscopy done in duplicates. Panels a-d show parental HEK293, e-h HEK293 cells overexpressing the wild type rat ABCC6 and i-l HEK293 cells overexpressing p.R389G mutant rat ABCC6. Green; rat ABCC6, red: plasma membrane marker Na-K ATPase, blue: Dapi staining. C) ATP efflux of the wild-type and R389G mutant Abcc6 overexpressed in HEK293 cells. D) PPi accumulation in the medium of HEK293 cells overexpressing the wild type or R389G rat ABCC6 after 24 hours incubation. Data in C and D depict the means +/-SE of representative experiments performed in octuple (n=8) in two independent experiments.

To further characterize the p.R389G sequence variant, we carried out biochemical tests using the rat ABCC6 p.R389G clone A2. ABCC6 mediates the efflux of ATP from the cytoplasm to the extracellular space (Jansen et al. 2014). The extruded ATP is rapidly cleaved to PPi and AMP by ENPP1, expressed in several cell types including HEK293 (Szeri et al. 2020). HEK293 cells overproducing rat ABCC6 p.R389G released similar amounts of ATP into the extracellular environment as wildtype rat ABCC6. Nor did the levels of PPi in the culture medium differ between both cell lines (Figure 3C,D). In conclusion, expression, subcellular localization and ATP release were unaltered by the p.R389G variant, indicating this mutation does not directly alter ABCC6 activity. These data support our hypothesis that ABCC6 interacts with other protein(s), which might play a decisive role in the development of PXE when ABCC6 harbors the p.R391G pathogenic variant.

## Discussion

PXE displays a remarkable though largely unexplained clinical and molecular heterogeneity. As IP of certain disease-causing alleles can be a potential mechanism to contribute to this heterogeienity, we sought to determine whether *ABCC6* IP pathogenic variants could be found in a large European cohort of PXE patients.

To this end, we used a recently created tool, with stringent exclusion criteria to avoid artefacts (Mikó et al. 2021). We applied correction for overrepresented homozygous pathogenic variants and therefore, only 52% of the initial alleles passed the filtering criteria. As a result of enrichment calculations, we found five variants to be more frequent in the reference population than in the patient cohort, thus we claim that these are non-pathogenic benign variants although identified as pathogenic previously. Other variants (p.G1327R, p.A950T, p.G1327E, p.A452D, p.E699K, p.R1339H) were enriched in the patient cohort, but not significantly. The pathogenicity of these variants was assessed recently by the Sherloc scoring system and the p.R1339H was found to be pathogenic while the others were classified as likely benign and of unknown significance (Verschuere et al. 2020). However, the population-genetic approach used herein requires larger well-characterized cohorts to determine their pathogenicity and penetrance.

We identified two incompletely penetrant variants: p.V787I and p.R391G, with a calculated 6.5% and 1.98% penetrance, respectively. This suggests, that only 6.5% and ∼2% of the carriers bearing another LOF variant on the second allele develop PXE. Similar low penetrant variants have been reported previously in several other Mendelian disorders (Minikel et al. 2016; Mikó et al. 2021).

Nevertheless, it should be emphasized that the p.V787I was reported previously in only one patient (Pfendner et al. 2007) and is present solely in two patients of our cohort, both originating from France. Due to the small number of patients harboring this pathogenic variant it was impossible to compare the severity of their phenotype to the biallelic LOF pathogenic variant phenotype. The V787 residue was previously shown to be evolutionarily conserved (Ran, Zheng, and Thibodeau 2018). Of note, based on protein glycosylation and expression studies it has also been suggested recently that the p.V787I ABCC6 has a similar cellular localization as the wild-type ABCC6 (Ran, Zheng, and Thibodeau 2018). Functional studies of the variant will be required once its IP will be further confirmed.

To assess the clinical symptoms of the patients with one p.R391G pathogenic variant we made use of the Phenodex scoring (Pfendner et al. 2007). Though the Phenodex scoring is somewhat debated, it is the most widely used tool for clinical evaluation of PXE. Although the number of patients with p.R391G was relatively small (n=17), our data showed that, when penetrant, it results in a phenotype as severe as that caused by the *ABCC6* LOF variants. This also suggests, that in rare cases (AF_in patients_^2^ = ∼1/2500 patients) patients with homozygous p.R391G pathogenic variants should also be detected amongst PXE patients. This was indeed recently reported for one patient (Boraldi et al. 2020). However, it should be noted that this patient also suffered from beta-thalassemia, which was previously shown to lead to a PXE phenocopy in older patients (Hamlin et al. 2003).

There is a further line of evidence, supporting the typical phenotype caused by the p.R391G variant. Recessive LOF pathogenic variants of *ENPP1* lead to generalized arterial calcification of infancy (GACI), a severe form of ectopic calcification, often leading to perinatal lethality (Ferreira et al. 2021). A strong relationship between PXE and GACI is supported by the reports of several patients with GACI phenotype and bi-allelic pathogenic variants in *ABCC6* in the absence of *ENPP1* variants (Nitschke et al. 2012). It should be noted that there were GACI patients diagnosed in different centers with p.R391G ABCC6 (Li et al. 2014; Nitschke et al. 2012). Although this variant has an AF of 0.78% in the general population, it is very unlikely that among the 34 unrelated *ABCC6-*related GACI patients, 3 patients would carry the p.(R391G) variant only by chance. This further confirms that in the appropriate context this IP pathogenic variant has full expressivity.

The low penetrance of p.R391G also suggests that in the gnomAD database containing genomic data from healthy individuals homozygous p.R391G variants are expected at a theoretical frequency of 6.1/100,000 since the AF is 0.78% in that population. Indeed, there are 2 homozygous healthy individuals (both >40years) among the 63124 European individuals in the gnomAD database, which is not different from the expected frequency (p=0.58). Furthermore, we have recently found an individual homozygous for p.R391G without PXE-related phenotype. However, these observations should be taken with caution since it is not completely excluded that patients with non-congenital, late-onset diseases may also be included in gnomAD (Karczewski et al. 2020). Additionally, it has been shown recently, that in some rare cases a very mild, eye-restricted form of PXE can develop in the elderly harboring the p.R391G pathogenic variant (Issa, Tysoe, and Caswell 2020), which most probably remains undetected in most of the cases.

Trying to find an explanation for the IP at the molecular level we hypothesized that the p.R391G variant becomes a classical disease-causing pathogenic variant only if another intramolecular variant of ABCC6 is co-occurring. This hypothesis was further strengthened by the homology models indicating potential interactions between the residue and other residues of the protein (Issa, Tysoe, and Caswell 2020; Szeri et al. 2021). However, when testing our PXE patient database for co-occurring sequence variants we did not find any *ABCC6* SNPs consistently present in *cis*. This ruled out the hypothesis of an intra-protein interaction leading to the development of PXE or GACI in the presence of the p.R391G pathogenic variant. Another explanation for our findings could be an interaction of the ABCC6 proteins expressed from the two different alleles. The interaction would then be abolished by the p.R391G pathogenic variant only in the presence of a specific pathogenic variant on the other allele. Such an intermolecular mechanism leading to IP has been reported in the case of the *NPHS2* gene (Tory et al. 2014). However, for the ABCC6 protein no such interaction or any indication of its di- or multimerization has ever been reported. Furthermore, we have not found any specific pathogenic variant pattern on the 2^nd^ allele (in *trans*), further disproving this scenario.

Ruling out these potential mechanisms, we hypothesized that IP of the p.R391G pathogenic variant can arise if ABCC6 is participating in a protein-protein interaction, that is crucial for the correct localization/function of ABCC6. Such an interaction partner has not yet been identified for ABCC6. However, in the ABCC gene family two proteins, SUR1 and SUR2, are ion channel regulators physically interacting with other membrane proteins (Ding et al. 2019; Foster and Coetzee 2016). It has also been shown, that CFTR interacts with several other proteins (Haggie et al. 2006; Ko et al. 2004). According to our hypothesis, the interaction between ABCC6 and the unidentified partner protein takes place through the R391 residue, which is in a highly conserved amino acid stretch, strengthening our hypothesis on its critical role (Figure 2). We hypothesize, that this interaction with the unknown protein would be ablated by the presence of the G391 variant only in the case when the interacting partner also has a rare sequence variant, unable to interact with G391 but still functional with the R391 residue. According to this theory, for the elimination of the interaction between the ABCC6 p.R391G and the other protein, both alleles of the interacting protein should be mutated, which would explain the very low penetrance of the p.R391G variant.

It should be emphasized that the putative interaction between ABCC6 and the yet unknown partner may occur at various levels. This interaction might take place temporarily during protein maturation or trafficking, could affect the localization, stability, turnover, or the plasma membrane retention of ABCC6. Last but not least, it may as well directly affect the primary physiological function of ABCC6, mediating cellular ATP efflux. Interestingly, an important proportion of ABCC6 missense variants are localization mutants, which retain their enzymatic activity if their plasma membrane localization is rescued. Hypothesizing that the interaction partner of ABCC6 involved in the p.R391G pathogenesis is related to the maturation and localization of ABCC6 might even give hints about potential therapies with chemical chaperones, e.g. 4-phenyl-butyrate (4-PBA), an FDA approved drug. Indeed, 4-PBA was already shown to rescue some mislocalized ABCC6 mutants retained in the ER (Saux et al. 2011; Pomozi et al. 2017).

ABCC6 is a transmembrane protein localized in the basolateral membrane (Pomozi et al. 2013), expressed mainly in hepatocytes, less in kidney and the gastrointestinal epithelial cells due to its transcriptional regulation governed by HNF4*α* (de Boussac et al. 2010; Arányi et al. 2005), but virtually absent from HEK293 cells. The direct transport of various substrates (e.g. LTC_4_) by ABCC6 was initially observed but could not be confirmed later. It has been shown more recently that the protein promotes ATP release from hepatocytes to the bloodstream (Jansen et al. 2014; Jansen et al. 2013), providing an indisputable explanation for the vast majority of the pathologic findings related to PXE. Importantly, the source of the extensive phenotypic heterogeneity observed in PXE has not yet been clarified unambiguously. Based on the IP variant identified here we propose that at least some of the heterogeneity can be explained by hypothesizing that ABCC6 requires a yet unidentified factor to exert its physiologic role. Our *in vitro* results indicate that a simultaneous (pathogenic) variant of this putative interaction partner is necessary to abolish the function of the p.R391G variant. We assume, that in the commercially available HEK293 cell line the unknown interacting partner is present in homozygous wild type or heterozygous form as the function of the variant was not altered.

Collectively, our data have important clinical relevance for genetic counseling. First, the p.R391G pathogenic variant can cause either PXE or GACI. Second, the expected clinical phenotype is not different from any other known pathogenic variants leading to these diseases. Third, and most importantly, based on our study this pathogenic variant has low penetrance, which probably relies on sequence variants in other gene(s), thus the co-occurrence of p.R391G dependent disease in siblings is probably much less frequent than expected. Along these lines it seems reasonable to consider pathogenic variants of low or very low penetrance as risk factors, in the general sense. The identification of the mechanism underlying penetrance will be crucial to provide correct preconceptional counseling in couples where one or both partners carry the p.R391G allele.

In conclusion, our results on the p.R391G pathogenic variant are leading to a new hypothesis on the molecular mechanism of ABCC6 strongly suggesting the necessity of an interacting partner. Our experimental data opens new avenues in PXE research for the identification of interacting partner(s), which might become important biomarker(s) and pharmacological target(s) in PXE or other ectopic calcification disorders.

## Methods (for details see Supplementary methods section)

### Patients

All PXE patients signed an informed consent. The study was approved by the University Hospital of Angers, the Ethics Committee of Ghent University Hospital, and of the University of Modena and Reggio Emilia. The Declaration of Helsinki was followed. The US patients were recruited under the IRB of Thomas Jefferson University, PA and the Genetic Alliance IRB – protocol PXE #001. Altogether 397 female and 191 male patients were included in the European cohort. The US cohort was composed of 478 patients. Patients were clinically diagnosed as described earlier (Vanakker et al. 2008; Pfendner et al. 2007; Boraldi et al. 2021) and mutation detection was performed as described earlier (and Supplementary methods).

### Bioinformatic analysis

Bioinformatic analysis was performed by the population-genetic based algorithm as described elsewhere (Mikó et al. 2021). Briefly, the unified patient cohorts underwent allelic sequence variant enrichment analysis relative to the European non-Finnish population of the gnomAD database. Significantly enriched alleles were tested for IP. The allele frequency of each significantly enriched (Fisher-exact test) variant was compared to the allele frequencies of fully penetrant complete loss-of-function (LOF) sequence variants. We considered the variant incompletely penetrant if the result of the comparison was lower than 30%, and significant after Bonferroni correction for multiple testing.

### Generation of mutant cell lines

Site-specific mutations were introduced into in pEntr223-rAbcc6 plasmid (Jansen et al. 2013) by Uracil excision-based (USER) cloning (Szeri et al. 2021).

### Immunoblot and Subcellular localization analyses of rat ABCC6

Rat ABCC6 was detected with the polyclonal K14 rabbit anti-rat ABCC6 antibody.

### Real-time ATP efflux assay and Quantification of PP_i_ levels in the medium of cells

Real-time ATP efflux assay was carried out and PP_i_ concentration of the medium samples were determined as described previously (Szeri et al. 2020).

### Statistical analyses

Data were analyzed using Prism 8.4.2 (GraphPad Software Inc.)

## Data Availability

All the data obtained are included in the manuscript.

## Acknowledgements

Authors gratefully acknowledge the collaboration of people affected by PXE and the support of PXE International and other patient’s associations. This work is a result of a collaboration in the frame of the EuroSoftCalcNet COST Action (CA16-115). This work was supported by the National Research, Development and Innovation Office [NKFIH, FK131946], U.S Department of State [Fulbright Visiting Scholar Program], Hungarian Academy of Sciences [Bolyai Janos Fellowship BO/00730/19/8, Mobility grant] and by the UNKP-2020 New National Excellence Program of the Ministry for Innovation and Technology from the source of the NKFIH to FS. Further funding for this work was provided by PXE International and the NIH NIAMS Grant R01AR072695 (KvdW), PXE Italia Odv (DQ). SV is a PhD Fellow supported by a Methusalem grant from the Ghent University - Belgium (BOF08/01M01108). OMV is a Senior Clinical Investigator of the Research Foundation Flanders (FWO) - Belgium. MTA-SE Lendulet Research Grant (LP2015-11/2015) of the Hungarian Academy of Sciences, the KH125566, K135798 grants to KT and the Ministry of Human Capacities in Hungary in the frame of Institutional Excellence Program for Higher Education. Diagnosis and management of PXE DNA samples occurred at the PXE Reference Center (MAGEC Nord), Angers University Hospital, Angers, France, as a part of the French PXE cohort (ClinicalTrials.gov Identifier: NCT01446380). TA is a beneficiary of the NKFIH K132695 grant.

