## Supplemental Methods for "The pathogenic p.R391G ABCC6 displays incomplete penetrance implying the necessity of an interacting partner for the development of pseudoxanthoma elasticum"

***Mutation detection***

Mutation detection was performed as described earlier. To distinguish pseudogenes from the *ABCC6* gene and determine large deletions (Pulkkinen et al. 2001; Pfendner et al. 2007; Symmons, Váradi, and Arányi 2008). Briefly, genomic DNA was isolated from whole blood (QIAamp blood kit, Qiagen®) according to the manufacturer’s protocol. Molecular analysis of the coding sequence and the intron-exon boundaries of the *ABCC6* gene was performed as previously described (Vanakker et al. 2008; Hosen et al. 2015). C-notations are based on NM_001171.5 and p-notations on NP_001162.4. MLPA analysis of the *ABCC6* gene was performed to evaluate the presence of larger deletions or insertions by using commercially available SALSA MLPA kit PO92-B3 (MRC-Holland) and according to the manufacturer’s recommendations ([www.mlpa.com](http://www.mlpa.com)).

***Cell culturing and generation of mutant cell lines***

HEK293 cells were cultured at 37^o^C and 5% CO_2_ in humidifying conditions in HyClone DMEM (GE) completed with 100 units pen/strep per ml (Gibco) and 5% FBS. Site-specific mutations were introduced into in pEntr223-rAbcc6 plasmid (Jansen et al. 2013) by Uracil excision-based (USER) cloning (Geu-Flores et al. 2007) with the following reverse and forward primers: ACCTTTCCGUACACCAGGCCAGTGATGGC and ACGGAAAGGUCCTGGTCCTGTCCAGTGGTTCCA, respectively. The cDNA encoding the pEnter223-rAbcc6 R389G mutant were sequenced and subsequently subcloned into a Gateway compatible pQCXIP expression vector and were transfected into HEK293 cells with calcium phosphate precipitation method (Szeri et al. 2021; Szeri, Niaziorimi, et al. 2020). The transfected cells were selected in completed DMEM medium also containing 2 µM puromycin. After puromycin selection the expression of the R389G rat ABCC6 in isolated cell clones was confirmed by immunoblot analysis and compared to that of the wild-type rat ABCC6 cell line. For the functional assays HEK293 cell lines were seeded in poly-D-lysine-coated 96-well plates in 100µl of completed DMEM, and experiments were conducted with wells using cells forming confluent monolayers (Szeri et al. 2021; Szeri, Niaziorimi, et al. 2020).

### Immunoblot and analysis of rat ABCC6

Cell lysates were prepared in lysis buffer (10 mM KCl, 10 mM Tris-HCl and 1.5 mM MgCl_2_, pH 7.4) supplemented with protease inhibitors (Roche). Samples containing 5 µg of total protein determined by BCA assay (Pierce™ BCA Protein Assay Kit, Thermo Scientific) were separated on 7.5% SDS-polyacrylamide gels (Bio-Rad) and transferred to a PVDF membrane with a semi-dry system (Bio-Rad). Wild-type and mutant rat ABCC6 were detected with the polyclonal K14 rabbit anti-rat ABCC6 antibody diluted by 1:3000 (kind gift of Dr. Bruno Stieger and HRP-conjugated donkey anti-rabbit secondary antibody (SA1200 Fisher Scientific) in a 1:5000 dilution. The signal was visualized by ECL (Pierce Western blotting substrate, Thermo Scientific).

### Subcellular localization of rat ABCC6 in HEK293 cells

Rat ABCC6 was detected as described previously(Szeri, Niaziorimi, et al. 2020). In short, HEK293 cells were seeded and grown for 2 days on ibi-Treat 1.5 µ-Slide 4 well chamber slides (80426, Ibidi) previously coated with poly-D-lysine. Cells were fixed in 4% PFA and subsequently in -20 ºC cold methanol for 5 min each. Samples were blocked with Protein Block solution (BioGenex) for 60 min. Coverslips were incubated with the polyclonal rabbit anti-rat ABCC6 antibody K14 diluted by 1:100 (kind gift of Dr. Bruno Stieger) and the mouse monoclonal anti-alpha 1 sodium potassium ATPase antibody (ab7671, Abcam) diluted by 1:250 for 60 min. Subsequently, samples were incubated with A488-conjugated anti-rabbit secondary antibody (A11008, Fisher Scientific) and A568 conjugated anti-mouse antibody (A11004, Fisher Scientific) both diluted by 1:1000 for 60 min. Samples were subsequently incubated with 300nM DAPI (40043, Biothium) for 5min to stain nuclei. The intracellular localization of the wild type and mutant rat ABCC6 were analysed by two point-scanning laser confocal microscope Nikon Eclipse T*i* equipped with a Nikon A1R+ at the Bioimaging Shared Resource of the Sidney Kimmel Cancer Center (NCI 5 P30 CA-56036).

### Quantification of PP_i_ levels in the medium of cells

Confluent HEK293 cells in 96-well plates were incubated in fresh medium for 24 hours. PP_i_ concentration of the medium samples were determined as described previously (Szeri, Lundkvist, et al. 2020). First, PPi was converted into ATP in an assay containing 50 mM HEPES pH 7.4, 80 µM MgCl_2_, 32 mU/ml ATP Sulfurylase (New England Biolabs) and 16 µM adenosine 5´-phosphosulfate (Sigma-Aldrich) by

incubating samples and standards at 37˚C for 30 min followed by inactivation of the enzyme at 90˚C for 10 minutes. In a consecutive step, ATP content was determined in a bioluminescent assay adding BacTiterGlo (Promega,) to samples and standards in a 1:1 ratio. PPi concentration of plasma samples was calculated with standard calibration. Values were corrected for the initial sample ATP concentrations.

### Real-time ATP efflux assay

Real-time ATP efflux assay were conducted as described previously (Szeri, Lundkvist, et al. 2020). HEK293 cells seeded in poly-D-lysine-coated black 96-well plates with an optically clear bottom were allowed to grow to confluence in 2 days in completed DMEM medium. At confluency the medium was removed and replaced by 50 μl efflux buffer, consisting of 11.5 mM HEPES (pH 7.4), 130 mM NaCl, 5 mM MgCl_2_, 1.5 mM CaCl_2_ and 11.5 mM glucose. Cells were incubated for 1 hour at 27 ºC in efflux buffer. Next, 50 µl efflux buffer containing 10% BactiterGlo (Promega) reactant previously dissolved in efflux buffer according to the instructions of the manufacturer was added to each well. Bioluminescence was subsequently determined in real time in a Flex Station 3 microplate reader (Molecular Devices). The real-time ATP efflux assay was run at 27 ºC for the first 1 hour and then at 37 ºC for 2 hours. The initial low temperature allowed the endogenous ectonucleotidases to metabolize the excess ATP generated by medium-change initiated sheer-stress resulting to an ABCC6-independent ATP efflux, that otherwise gave a significant background in the experiments.

### Statistical analyses

Data were analyzed using Prism 8.4.2 (GraphPad Software Inc.). Correlation between age and Phenodex score was assessed by linear regression with 95% confidence intervals. Two-tailed t-test was applied to test significance of pyrophosphate concentrations of HEK293 cells overexpressing the wild type or the p.(R391G) variant. Significance was accepted at p<0.05.
